# Human bronchopulmonary disposition and plasma pharmacokinetics of oral bemnifosbuvir (AT-527), an experimental guanosine nucleotide prodrug for COVID-19

**DOI:** 10.1101/2024.01.03.24300783

**Authors:** Xiao-Jian Zhou, Arantxa Horga, Adeep Puri, Lee Winchester, Maureen Montrond, Keith Pietropaolo, Bruce Belanger, Courtney V. Fletcher, Janet Hammond

## Abstract

Bemnifosbuvir (BEM, AT-527) is a novel oral guanosine nucleotide antiviral drug for the treatment of patients with COVID-19. Direct assessment of drug disposition in the lungs, via bronchoalveolar lavage, is necessary to ensure antiviral drug levels at the primary site of SARS-CoV-2 infection are achieved. We conducted a Phase 1 study in healthy subjects to assess the bronchopulmonary pharmacokinetics, safety, and tolerability of repeated doses of BEM. A total of 24 subjects were assigned to receive twice-daily (BID) BEM at doses of 275, 550, or 825 mg for up to 3.5 days. AT-511, the free base of BEM, was largely eliminated from the plasma within 6 h post dose in all dosing groups. Antiviral drug levels of BEM were consistently achieved in the lungs with BEM 550 mg BID. The mean level of the guanosine nucleoside metabolite AT-273, the surrogate of the active triphosphate metabolite of the drug, measured in the epithelial lining fluid of the lungs was 0.62 µM at 4–5 h post dose. This exceeded the target *in vitro* 90% effective concentration (EC_90_) of 0.5 µM for antiviral drug exposure against SARS-CoV-2 replication in human airway epithelial cells. BEM was well tolerated across all doses tested, and most treatment-emergent adverse events reported were mild in severity and resolved. The favorable pharmacokinetics and safety profile of BEM demonstrates its potential as an oral antiviral treatment for COVID-19, with 550 mg BEM BID currently under further clinical evaluation in patients with COVID-19.

## INTRODUCTION

The COVID-19 pandemic caused by SARS-CoV-2 continues to be a worldwide public-health concern, with more than 6.9 million confirmed deaths reported globally according to the World Health Organization (https://www.who.int/emergencies/diseases/novel-coronavirus-2019). SARS-CoV-2 infects the respiratory tract and, although the nasopharynx is an initial portal of entry and site of infection commonly used in testing, it is infection of the lower airway and lungs that results in substantial mortality (1–4). Effective drug delivery to the critical site of infection and prevention of disease progression is therefore important for the treatment of COVID-19. Currently, remdesivir and nirmatrelvir/ritonavir are the only antiviral drugs approved by the FDA for the treatment of COVID-19, and molnupiravir has received emergency use authorization by the FDA (https://www.fda.gov/news-events/press-announcements/fda-approves-first-treatment-covid-19; https://www.fda.gov/news-events/press-announcements/fda-approves-first-oral-antiviral-treatment-covid-19-adults; https://www.fda.gov/news-events/press-announcements/coronavirus-covid-19-update-fda-authorizes-additional-oral-antiviral-treatment-covid-19-certain). However, remdesivir is not suitable for oral delivery due to its rapid clearance, and is limited to use in a hospital or alike setting due to intravenous infusion as the only route of administration (5–7). Many drug–drug interactions between nirmatrelvir/ritonavir and concomitant medications have been identified in patients with COVID-19 (8–11). Furthermore, uncertainties regarding the mutagenicity of molnupiravir towards mammalian host DNA have been reported, as well as evidence of molnupiravir-induced mutations in the virus sequence which could potentially lead to new variants of SARS-CoV-2 (12–16). Although the pharmacokinetics (PK) of these antiviral drugs against

SARS-CoV-2 have previously been studied (17–19), there is a lack of clinical data regarding the attainment of antiviral drug levels in the lungs, the site of SARS-CoV-2 replication, which can be measured by bronchoalveolar lavage (BAL). Despite the high viral load of SARS-CoV-2 in the lung tissue, data suggest that repurposed antiviral drugs used to treat COVID-19 may not reach an effective concentration in the lungs necessary to inhibit viral replication (20, 21). Hence, there is an unmet need for direct-acting antivirals that can be widely administered in an outpatient setting and are demonstrated to effectively target the primary site of infection.

Bemnifosbuvir (BEM, AT-527) is an orally available guanosine nucleotide prodrug that has demonstrated potent antiviral activity against hepatitis C virus (HCV) and SARS-CoV-2 (22–24). BEM is the hemisulfate salt of AT-511, a novel phosphoramidate protide that is converted after multistep activation to the active 5ill triphosphate metabolite AT-9010, which is formed only after intracellular delivery of the prodrug (**Figure 1**) (23, 24). AT-9010 selectively inhibits viral replication by a dual mechanism of action, simultaneously targeting the RNA-dependent RNA polymerase (RdRp) and nidovirus RdRp-associated nucleotidyl transferase (NiRAN) domains of the SARS-CoV-2 RNA polymerase, resulting in potent inhibition of viral RNA synthesis (25). Dephosphorylation of AT-9010 results in the formation of the guanosine nucleoside metabolite AT-273, which is regarded as a surrogate plasma marker for intracellular concentrations of AT-9010 (24).

**Figure 1.**
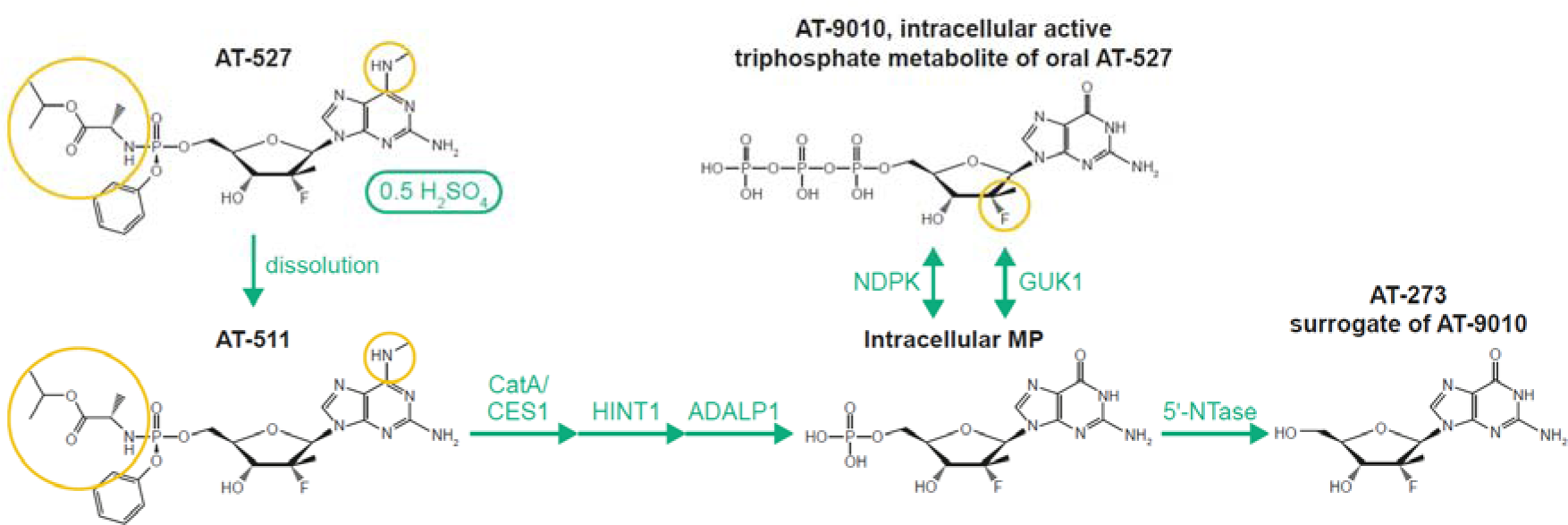
Activation pathway of BEM (AT-527). Circles indicate BEM structural features critical to its potent antiviral activity/lack of toxicity. 5⍰-NTase, 5⍰-nucleotidase; ADALP1, adenosine deaminase-like protein 1; BEM, bemnifosbuvir; CatA, cathepsin A; CES1, carboxylesterase 1; GUK1, guanylate kinase 1; HINT1, histidine triad nucleotide binding protein 1; MP, monophosphate; NDPK, nucleoside-diphosphate kinase.

Once daily (QD) dosing of BEM up to 550 mg in healthy volunteers and HCV-infected patients has previously been assessed in two clinical trials (ClinicalTrials registration no. NCT03219957; ClinicalTrials registration no. NCT04019717). Although the human PK profile reported for BEM in HCV-infected patents is also likely to be observed in patients with COVID-19, epithelial lining fluid (ELF) and the alveolar macrophages (AM) that reside in the ELF are important sites of infection in COVID-19, and therefore information about drug concentrations in the extracellular and intracellular compartments of the lung is essential for development of a drug targeting lower respiratory tract infections.

This Phase 1 study was designed to evaluate antiviral drug levels of BEM in the lungs of healthy volunteers using BAL to demonstrate attainment of antiviral drug levels at the primary site of SARS-CoV-2 infection, and to assess the safety and tolerability of BEM when given as repeated oral doses. Dosing of BEM was informed by the previously predicted lung exposure of its active triphosphate metabolite AT-9010, using plasma levels of its surrogate nucleoside metabolite AT-273, to exceed the *in vitro* 90% effective concentration (EC_90_) of 0.5 μM BEM for inhibition of SARS-CoV-2 replication (24, 26). BEM 550 mg twice daily (BID) was expected to produce sustained lung exposure to AT-9010 greater than the concentration required to inhibit SARS-CoV-2 replication in human airway epithelial (HAE) cells *in vitro* (24). Although the 550 mg BID regimen was believed to be optimal, a lower and higher dose were also evaluated so that a dose-response relationship could be established.

## RESULTS

### Subject demographics and baseline characteristics

A total of 24 healthy subjects entered and completed the trial, and no subjects were withdrawn. All enrolled subjects were included in the safety population, PK concentration population, and PK parameter population. Demographics and baseline characteristics are summarized in **Table 1**. The groups across dosing levels were generally comparable, with sex being the only notable difference across dosing levels: one woman (12.5%) received 275 mg BEM BID, while an equal number of men and women (four each per dose) received the other dose levels (550 mg and 825 mg BID). There were no other notable demographic differences between groups. No subjects identified their ethnicity as Hispanic or Latino.

**Table 1.** Baseline characteristics of study subjects.

| Variable | 275 mg BEM<br>BID<br>n=8 | 550 mg BEM<br>BID<br>n=8 | 825 mg BEM<br>BID<br>n=8 | All subjects<br>N=24 |
| --- | --- | --- | --- | --- |
| Mean age, years (range) | 42.3 (23–64) | 44.0 (27–62) | 40.6 (21–64) | 42.3 (21–64) |
| Female, n (%) | 1 (12.5) | 4 (50.0) | 4 (50.0) | 9 (37.5) |
| Male, n (%) | 7 (87.5) | 4 (50.0) | 4 (50.0) | 15 (62.5) |
| Race, n (%) |  |  |  |  |
| Black or African American | 0 | 0 | 2 (25.0) | 2 (8.3) |
| White | 8 (100.0) | 8 (100.0) | 5 (62.5) | 21 (87.5) |
| Other | 0 | 0 | 1 (12.5) | 1 (4.2) |
| Mean weight, kg (range) | 79.1 (67.2–96.4) | 72.2 (56.4–94.5) | 69.5 (56.6–83.3) | 73.6 (56.4–96.4) |
| Mean BMI, kg/m<sup>2</sup> (range) | 24.4 (20.1–27.9) | 25.2 (22.8–27.9) | 23.8 (20.8–26.0) | 24.5 (20.1–27.9) |

BEM, bemnifosbuvir; BID, twice daily; BMI, body mass index.

### Safety and tolerability

BEM was well tolerated across all doses, and no serious adverse events (SAEs) or adverse events (AEs) leading to discontinuation were observed during the study. 11 subjects (45.8%) reported at least one treatment-emergent adverse event (TEAE) after the start of BEM dosing (17 reported in total). Of the 11 subjects reporting a TEAE, two subjects (25.0%) had received 275 mg, three (37.5%) had received 550 mg, and six (75.0%) had received 825 mg BEM BID. The most frequently reported TEAEs were nausea (n=2; 825 mg BEM BID), esophageal mucosa erythema (n=2; 825 mg BEM BID), and oropharyngeal pain (n=2; one each at 275 mg and 825 mg BEM BID). All other TEAEs were reported in one subject each (dizziness, headache, lethargy, paresthesia, tremor, chest pain, pyrexia, vessel puncture site bruise, increased C-reactive protein [CRP] and leukocyte count, and back pain). The severity of most TEAEs was considered to be mild, except for three that were Grade 2 in severity (headache, chest pain, and pyrexia), each reported by one subject per dosing level. Of the 17 TEAEs reported, two (both mild nausea) were considered to be possibly related to the study drug, both after the second dose of 825 mg BEM BID and lasting 1–2 h. Three TEAEs had unknown outcomes and were reported during subjects’ final follow-up visit (lethargy and two instances of esophageal mucosa erythema), and all other TEAEs resolved. One subject who received 825 mg BEM BID reported a fever after the final dose, approximately 4 h after bronchoscopy, although this was likely a side effect of the midazolam sedation medication used during bronchoscopy. The fever coincided with a high leukocyte count and raised CRP level, which was considered clinically significant; however, the leukocyte count was within the reference range at all other timepoints and the CRP level returned to normal by the time of the subject’s follow-up visit.

### Plasma PK

Mean (± standard deviation [SD]) plasma concentration–time profiles for AT-511 and the guanosine nucleoside metabolite AT-273 (surrogate of the intracellular active triphosphate AT-9010) are presented in **Figure 2**. The mean AT-511 plasma concentration–time curves were similar in shape after the last dose of 275 mg, 550 mg, and 825 mg BEM BID. Plasma PK parameters of AT-511 and AT-273 are summarized in **Table 2**. AT-511 was rapidly absorbed in plasma after oral dosing, with the rate of absorption unaffected by dose: median time to maximum plasma concentration (t_max_) was 0.5 h after all doses of BEM. AT-511 was largely eliminated from plasma within 6 h post dose with a short half-life of approximately 1 h in all dosing groups, and the concentration–time profile was consistent with first-order PK. Mean maximum plasma concentration (C_max_) and area under plasma concentration–time curve during a dosing interval (AUC_τ_) of AT-511 increased more than proportionally at the lower dosing levels of 275 mg and 550 mg BEM BID, and less than proportionally at the higher dosing levels of 550 mg and 825 mg BEM BID.

**Figure 2.**
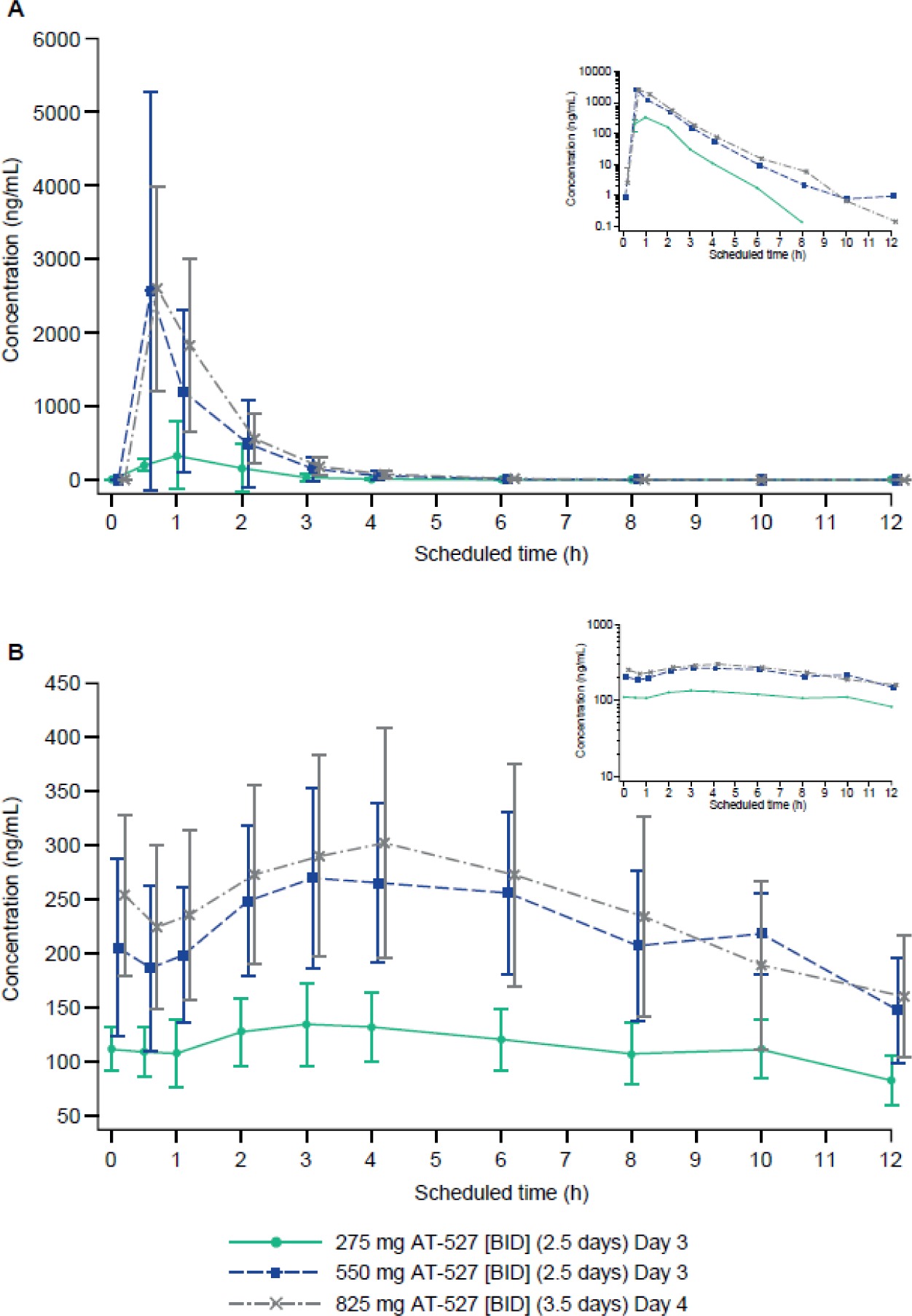
Mean (± SD) plasma concentration–time profiles of AT-511 (A) and AT-273 (B) on the last dosing day up to 12 h post dose of 275 mg, 550 mg, and 825 mg BEM dosage regimens (inserts: semi-log plots with no SD for clarity). BEM, bemnifosbuvir; BID, twice daily; SD, standard deviation. ‘0 h’ is pre dose (morning), ‘4 h’ is 4 h or pre BAL (4–5 h, as applicable), and ‘10 h’ is pre BAL (11–12 h). Values below the limit of quantification are imputed to 0. Panel A: At each dose level, n=8 except at 10 h (n=4).

**Table 2.**
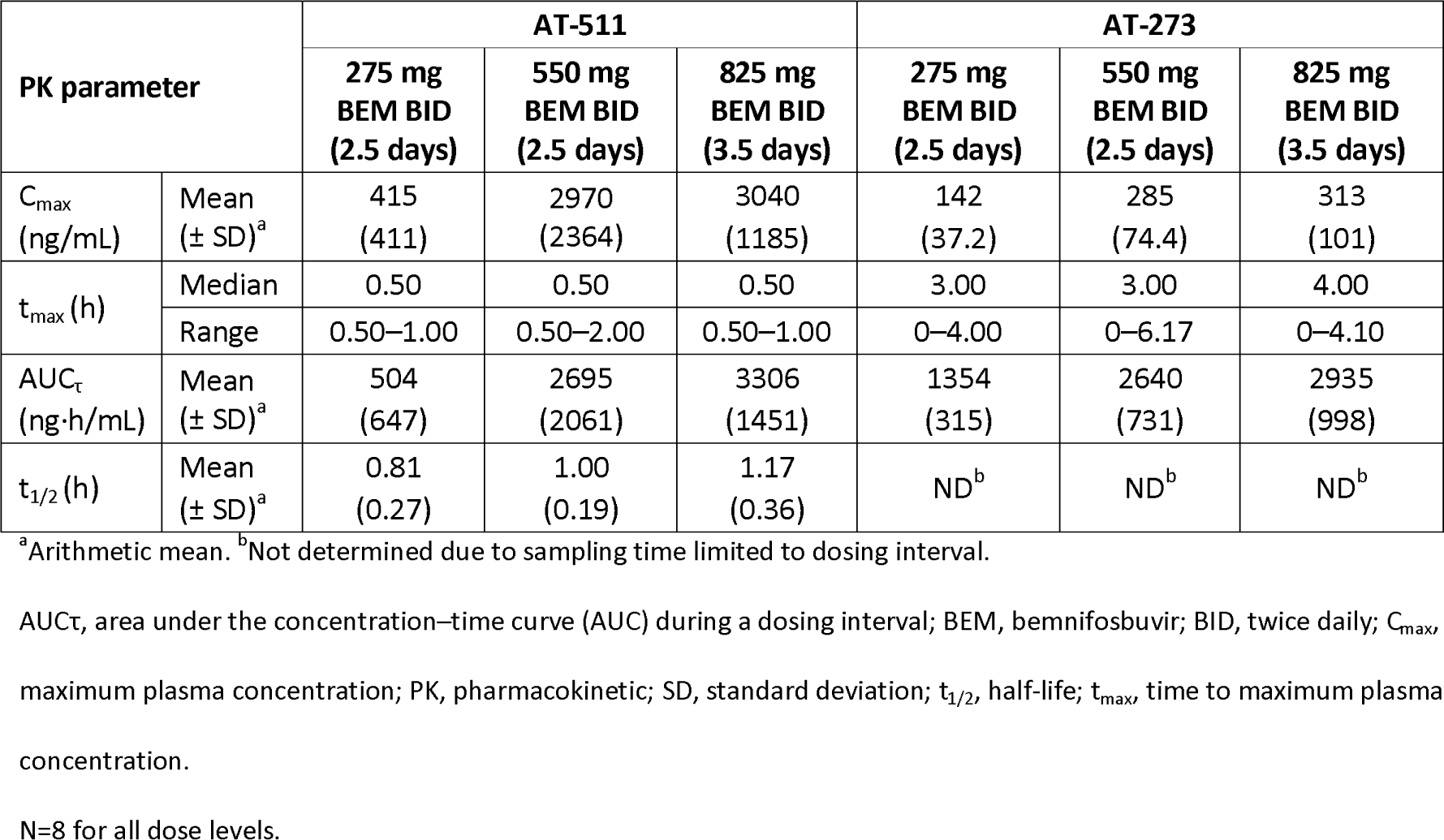
Summary of repeat dose plasma PK parameters of AT-511 and AT-273: PK parameter population.

After BEM BID, median t_max_ of AT-273 was 3–4 h across dose levels. Mean C_max_ and AUC_τ_ of AT-273 increased proportionally with dose. The mean (± SD) trough plasma concentrations (C_trough_) of AT-273 at 12 h after the final dose of 275 mg, 550 mg, and 825 mg BEM BID were 92.1 (27.2), 168.0 (51.8), and 144.7 (47.6) ng/mL, respectively. Plasma concentrations of AT-273 were sustained without much fluctuation and consistently exceeded the target level of 150 ng/mL (0.5 μM, EC_90_ of the drug for inhibiting SARS-CoV-2 replication in HAE cells *in vitro*) over the dose interval with BEM 550 mg BID (**Figure 2**).

### ELF and time-matched plasma concentrations

Mean (± SD) ELF and time-matched plasma concentrations of AT-273 are presented in **Table 3**. All AT-511 concentrations in ELF were below the limit of quantification (BLQ), except for one subject who had an AT-511 concentration of 10.5 ng/mL in ELF on Day 4, at the 4–5 h BAL timepoint. Concentrations of AT-273 were quantifiable in BAL fluid supernatant (BALF) at the two highest dose levels of BEM (550 mg and 825 mg BID), regardless of BAL timepoint. After 275 mg BEM BID, three of four subjects who had their BAL at 11–12 h post dose had quantifiable AT-273 in BALF, but all four of those who had their BAL at 4–5 h post dose did not. Overall, ELF AT-273 concentrations were higher among subjects who had their BAL procedure at 4–5 h post dose than in those who had BAL at 11–12 h post dose, approximately corresponding to the peak and trough plasma levels.

**Table 3.** Mean (± SD) comparative ELF vs plasma concentrations of AT-273.

| Dose | AT-273 concentration (ng/mL) | Sampling time (h) |  |
| --- | --- | --- | --- |
|  |  | Day 3 or 4<sup>a</sup><br>4–5 h | Day 3 or 4<sup>a</sup><br>11–12 h |
| 275 mg BEM BID (2.5 days) | Mean ELF (± SD) | 0 | 53.3 (44.2) |
| | Mean plasma (± SD) | 117 (21.4) | 92.1 (27.2) |
| | Ratio (± SD) | 0 | 0.72 (0.10)<sup>b</sup> |
| 550 mg BEM BID (2.5 days) | Mean ELF (± SD) | 185 (89.6) | 138 (76.0) |
| | Mean plasma (± SD) | 240 (39.4) | 168 (51.8) |
| | Ratio (± SD) | 0.76 (0.30) | 0.79 (0.18) |
| 825 mg BEM BID (3.5 days) | Mean ELF (± SD) | 197 (96.2) | 103 (24.0) |
| | Mean plasma (± SD) | 330 (110) | 145 (47.6) |
| | Ratio (± SD) | 0.58 (0.20) | 0.73 (0.09) |

<sup>a</sup>Last dosing day was on Day 3 for Groups 1 and 2 (275 and 550 mg BID) and on Day 4 for Group 3 (825 mg BID). <sup>b</sup>n=3.
BEM, bemnifosbuvir; BID, twice daily; ELF, epithelial lining fluid; SD, standard deviation.
0.5 μM equivalent to 150 ng/mL for AT-273. Half of each cohort had the BAL procedure 4–5 h after dosing, the other half at 11–12 h after dosing. At each dose level, n=8 except at 11–12 h (n=4). ELF concentrations BLQ were imputed to 0 before correcting the values for (plasma/BALF) urea ratio.

At the 550 mg BEM BID dose, mean ELF levels of AT-273 at the 4–5 h and 11–12 h BAL timepoints were 185 and 138 ng/mL (0.62 and 0.46 µM), respectively (exceeding or approaching the target level of 150 ng/mL or 0.5 µM). Similarly, at the 825 mg BEM BID dose, mean ELF levels of AT-273 at the 4–5 h and 11–12 h BAL timepoints were 197 and 103 ng/mL (0.66 and 0.34 µM), respectively. Mean ratios of ELF-to-plasma concentrations were generally below 1 (range of 0.58 to 0.78). There was a significant correlation between ELF and plasma levels of AT-273 (r^2^=0.6777, p<0.0001) (**Figure 3**).

**Figure 3.**
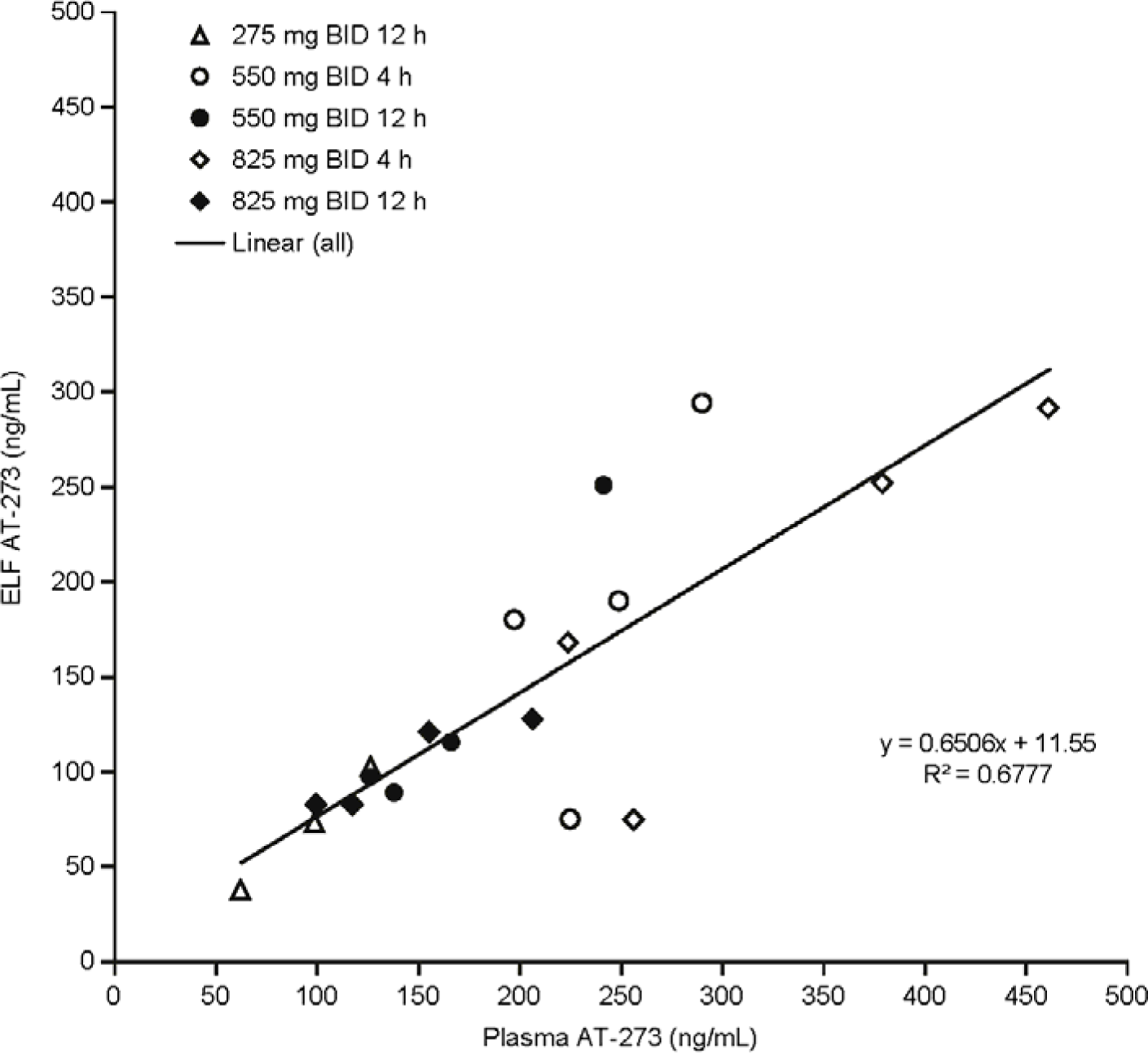
Correlation between plasma and ELF AT-273 levels. BID, twice daily; ELF, epithelial lining fluid. ELF AT-273 levels with 275 mg BID at the 4 h timepoint were BLQ and therefore excluded.

### AT-9010 concentrations

AM samples for the 275 mg and 550 mg BEM BID doses were left at room temperature without dry ice for 11 days (May 24 to June 4, 2021) during transfer, resulting in degradation of AT-9010 and making data uninterpretable. After 825 mg BEM BID, AT-9010 in AM was quantifiable (>lower limit of quantification [LLQ] at 3.34 fmol) in all but one subject who had their BAL at 11–12 h post dose. In the remaining subjects, AT-9010 concentrations in AM ranged from 0.79 to 5.37 nM.

## DISCUSSION

The PK of orally administered BEM has previously been reported in HCV-infected patients, and briefly described in healthy subjects and patients with COVID-19 (22, 26, 27); however, the delivery to the lungs of BEM or any other direct-acting antivirals against SARS-CoV-2 has not previously been studied. This study describes the first results of a Phase 1 study assessing the intrapulmonary disposition and plasma PK of BEM following various doses in healthy adults. BEM was well tolerated across all doses of 275, 550, and 825 mg BID tested in healthy volunteers, with no clinically significant physical examination findings, vital signs, or electrocardiogram (ECGs) observed. There were no deaths, non-fatal SAEs, nor other significant AEs or AEs leading to subject withdrawal from treatment during the study. All TEAEs reported were mild or moderate in severity. The overall rate of TEAEs reported increased with increasing doses of BEM, with 25% of subjects reporting mild nausea after 825 mg BEM BID, which was considered as possibly related to the study drug. Two clinically significant laboratory results were reported in one subject receiving 825 mg BEM BID (high leukocyte count and raised CRP) coinciding with a fever; however, these symptoms were considered likely a result of the midazolam sedation used during bronchoscopy. BEM was well tolerated at the 550 mg BID dose, with no gastrointestinal TEAEs reported. This is consistent with the favorable safety profile of 550 mg BEM previously observed in HCV-infected subjects (22), the administration of 550 mg BEM QD in combination with daclatasvir for up to 12 weeks (28), and 550 mg BID BEM for 5 days assessed in both hospitalized patients with moderate COVID-19 and non-hospitalized patients with mild-to-moderate COVID-19 (29, 30).

As designed, AT-511, a phosphoramidate protide, exhibited a very transient plasma profile with fast absorption followed by rapid multistep activation to the intracellular antiviral triphosphate AT-9010. The active triphosphate was then dephosphorylated to the guanosine nucleoside metabolite, AT-273, which entered general circulation. As AT-273 cannot efficiently be anabolized to its phosphorylated forms, circulating AT-273 is therefore considered a surrogate of AT-9010. Oral dosing of BEM 550 mg BID resulted in sustained plasma levels of AT-273, which consistently exceeded the target of 150 ng/mL or 0.5 µM, the *in vitro* EC_90_ of AT-511 for inhibiting SARS-CoV-2 replication in HAE cells.

The BAL samples were obtained at 4–5 h and 11–12 h post dose to capture the maximum and trough ELF levels of AT-273. Due to its rapid clearance, AT-511 was not expected to be frequently measurable at these timepoints in the BAL fractions. In fact, AT-511 concentrations in BAL samples were BLQ in all but one subject. By contrast, AT-273 concentrations were mostly quantifiable in BAL samples, especially with the 550 mg BID and 825 mg BID doses. The 275 mg BID dose, however, did not achieve sufficient AT-273 levels. BEM 550 mg BID achieved ELF levels of AT-273, surrogate of AT-9010, exceeding or approaching the target 150 ng/mL or 0.5 µM at 4–5 h and 11–12 h, providing direct evidence that BEM 550 mg BID produced antiviral drug exposure in the lungs and confirming previously predicted lung AT-9010 levels with this regimen (24). The 825 mg BID regimen also produced ELF AT-273 levels exceeding the target at 4–5 h, but increased less than dose- proportionally from the 550 mg BID dose. ELF and plasma levels of AT-273 were found to be significantly correlated, potentially allowing for prediction of ELF levels from plasma samples, which are more commonly available. This relationship, taken together with previous results showing that plasma levels of AT-273 rapidly increased to exceed the target 150 ng/mL within a few hours of the first dose of BEM 550 mg BID (26, 27), suggests that BEM produced antiviral drug exposure in the lungs shortly following initiation of treatment. Since direct-acting antivirals are most effective when administered during the early stage of SARS-CoV-2 infection (7), high antiviral drug levels in the lungs achieved early after administration of BEM 550 mg BID are of particular relevance to a better treatment outcome. This is supported by the findings that BEM 550 mg BID for 5 days reduced hospitalization rates in patients with COVID-19 by 71% compared with placebo (n=207) (ClinicalTrials registration no. NCT04889040) (30).

In summary, the results of this study provide important information on the PK disposition of oral BEM in the lungs of healthy subjects assessed by BAL, which is the primary site of SARS-CoV-2 infection. We have confirmed the formation of AT-9010, the active triphosphate metabolite of BEM, in alveolar macrophages and demonstrated that the dosing regimen of 550 mg BEM BID achieved the target antiviral drug exposure in the lungs of 0.5 µM EC_90_ against SARS-CoV-2 replication, with a mean AT-273 ELF concentration of 0.62 µM at BAL taken 4–5 h post dose, exceeding the target antiviral level. In addition to the favorable PK demonstrated, BEM was well tolerated, with no significant AEs reported. BEM is therefore an attractive antiviral treatment for COVID-19, and the oral route of administration allows for use in the outpatient setting. A global Phase 3 clinical trial (ClinicalTrials registration no. NCT05629962) is currently ongoing to evaluate the safety and efficacy of oral BEM 550 mg BID for the treatment of high-risk outpatients with COVID-19.

## MATERIALS AND METHODS

### Study design and subjects

This was a Phase 1, open-label study to assess the bronchopulmonary PK, safety, and tolerability of repeated doses of BEM in healthy subjects (ClinicalTrials.gov registration no. NCT04877769). Healthy adult subjects between the ages of 18 and 65 who met the inclusion and exclusion criteria were considered eligible for this study. All subjects were required to have an initial screening that included clinical history, physical examination, 12-lead ECG, vital signs, spirometry, and laboratory tests of blood and urine. A follow-up visit was scheduled for 7 days after the last dose. All subjects were required to weigh at least 50 kg and have a body mass index of between 18 and 29 kg/m^2^, inclusive. Subjects were also required to have a forced expiratory volume (FEV_1_) and forced vital capacity (FVC) ≥80% of predicted value and FEV_1_/FVC ratio >0.7 within 28 days before their first dose of trial medication. Exclusion criteria included clinically relevant abnormal medical history, physical findings, ECG, or laboratory values at the pre-trial screening assessment; positive test for SARS-CoV-2 or suspected exposure to the SARS-CoV-2 virus during the 14 days before screening; receipt of a vaccine against COVID-19 in the 14 days before the first dose; or scheduled to receive a dose of the vaccine 2 weeks after the last dose. Subjects could not have received a prescription medicine during the 28 days before the first dose, and no over- the-counter medicine or herbal or dietary supplements were permitted during the 7 days before the first dose. Subjects with positive tests for hepatitis B antigen, hepatitis C, or human immunodeficiency virus were excluded. Subjects were allocated to receive either 550 mg of BEM for 2.5 days (Group 1), 275 mg of BEM for 2.5 days (Group 2), or 825 mg BEM for 3.5 days (Group 3), up to seven doses in total. Each dose was administered orally as either one or three tablets and taken with approximately 240 mL water.

The study protocol was approved by the UK Medicines and Healthcare products Regulatory Agency and West Midlands – Edgbaston Research Ethics Committee before the trial began, and written informed consent was obtained from each subject.

### Blood sample collection for plasma PK and urea

Blood samples (3 mL/timepoint) were obtained to determine drug and urea concentrations. On the day of BAL procedure (Day 3 for Groups 1 and 2; Day 4 for Group 3), serial blood samples for assay of plasma drug levels were taken before the final dose and 0.5, 1, 2, 3, 4, 6, 8, and 12 h post dose. For all groups, blood samples were also taken immediately before the bronchoscopy (performed at 4–5 h or 11–12 h after final dose) for assay of BAL time-matched plasma drug and urea levels. Blood samples were collected and immediately placed on ice, and centrifuged at 1500 × *g* for 10 min at 4°C. After centrifugation, the plasma was divided into two aliquots and snap frozen on dry ice within 60 min of collection, then stored at -80°C until analyzed. Analysis of AT-511 and metabolites was performed by Altasciences Company, Inc (Laval, Quebec, Canada), and urea analysis was performed by MicroConstants, Inc (San Diego, CA, USA).

### Bronchoscopy and BAL

BAL samples were taken 4–5 h and 11–12 h after dosing on Day 3 (Groups 1 and 2) or Day 4 (Group 3). The subjects were prepared with 6% lidocaine gel for nasal local anesthesia and lubrication. Midazolam 1 mg/mL with a 0.9% saline flush was prescribed for sedation if needed. The saline for lavage was drawn up in four 50 mL aliquots for a total volume of 200 mL. Up to four or five BAL samples (BAL 1–5) were collected for each subject. BAL 1 was processed alone, while BAL 2–4 (or BAL 2–5, if a fifth sample was collected) were pooled together for processing. For each of the two resulting samples, 2 × 4 mL BAL fluid was sent on ice to the laboratory for cell count and differential, while the remaining BAL fluid was centrifuged at 400 × *g* for 5 min at 4°C to separate the BALF and pellet. From the BALF, 4 × 2 mL aliquots (two for urea and two for PK) were prepared in cryovials and frozen at -80°C. The pellets (containing AM) were resuspended in ice cold 70% methanol solution containing 5 mM dibromoacetic acid and frozen on dry ice or at -80°C. The BALF and AM samples were stored at -80°C until shipped, frozen and on dry ice. For the BALF samples, analysis of AT- 511 and AT-273 was performed by Altasciences Company, Inc (Laval, Quebec, Canada) and urea analysis was performed by MicroConstants, Inc (San Diego, CA, USA). For the AM samples, analysis of AT-9010 was performed by the University of Nebraska Medical Center (Omaha, NE, USA).

### AT-511 and AT-273 plasma and BAL assays

Concentrations of AT-511 and AT-273 measured in plasma and BAL supernatants were determined by liquid chromatography tandem mass spectrometry (LC-MS/MS) (Altasciences Company, Inc). BAL samples were collected and mixed with an anti-adsorptive agent solution prior to storage. Following deproteinization with cold acetonitrile and methanol (1:3), samples were subsequently diluted for injection. Chromatographic separation involved gradient elution on an XBridge C_18_ column (50 × 4.6 mm, 3.5 µm) using 10 mM ammonium bicarbonate and acetonitrile and methanol (1:1) as organic modifier. The LC system was coupled to a SCIEX API 5000 mass spectrometer operated in positive electrospray ionization mode monitoring the multiple reaction monitoring (MRM) transitions 582.2>376.2 (AT-511) and 300.1>152.2 (AT-273). Analytical ranges were between 1 (LLQ) and 1000 ng/mL for both analytes in plasma, 0.200 (LLQ) and 200 ng/mL for AT-511, and 0.400 (LLQ) and 400 ng/mL for AT-273 in BAL.

### AT-9010 assay

Concentrations of AT-9010 in AM and other cell populations isolated from the upper airway lavage fluid were determined by LC-MS/MS (University of Nebraska Medical Center). Briefly, AT-9010 was extracted from cell debris via anion-exchange solid-phase extraction (SPE) followed by dephosphorylation to AT-273 via acid phosphatase. AT-273 was desalted by a reversed-phase SPE cartridge and analyzed using LC-MS/MS. Chromatographic separation was carried out on a Synergi Polar-RP 100 × 2.00 mm column for the stationary phase, using 5% acetonitrile, 0.1% formic acid in water, with 0.1% formic acid in acetonitrile with gradient elution for the mobile phase. AT-273 was detected by an AB Sciex 6500 triple quadrupole mass spectrometer in positive ion mode monitoring MRM transition 300.0>152.1. The analytical range was from 3.34 (LLQ) to 3340 fmol/sample. A mean alveolar macrophage cell volume of 2.42 μL/10^6^ cells was used in calculating AT-9010 concentrations (31).

### Urea plasma and BAL assays

Plasma urea concentrations were measured using a colorimetric detection assay with an assay range from 2.5 (LLQ) to 50 mg/dL. BAL urea concentrations were quantified using an immunoassay with an assay range from 0.15 to 2.5 mg/dL using 150 μL of sample, and from 0.05 to 1 mg/dL using 300 μL of sample.

### Calculation of the volume of ELF and drug concentrations

Drug levels in lung ELF were calculated by correcting for urea levels in the BAL and the corresponding plasma samples as: Conc_ELF_ = Conc_BAL_ × (Urea_Plasma_/Urea_BAL_), where Conc_ELF_ and Conc_BAL_ are drug levels in the ELF and BAL, respectively, and Urea_Plasma_ and Urea_BAL_ are the plasma and BAL urea levels, respectively.

### PK analysis

Plasma PK parameters were derived from plasma concentration data, using non-compartmental analysis in WinNonLin v8.3. C_max_, T_max_, and C_trough_ were calculated directly from the concentration–time data. AUC_τ_ after the final dose was calculated using the trapezoidal method. The PK concentration data were summarized using the PK concentration population and PK parameters were summarized using the PK parameter population. The pooled BAL 2–4 samples (or BAL 2–5, where a fifth BAL sample was collected) were used for the ratios and summary statistics.

### Statistical analysis

All statistical analyses were carried out using SAS^®^ version 9.4. PK analysis was carried out using non-compartmental approaches in WinNonlin version 8.3.

### Laboratory and safety assessment

Safety was measured by clinical laboratory tests (hematology, biochemistry, coagulation, serum chemistry, and urinalysis), physical examination, vital signs, 12-lead ECGs, spirometry, SARS-CoV-2 testing, and monitoring of AEs. All AEs were coded using the Medical Dictionary for Regulatory Activities (MedDRA) v24.1. Concomitant medications were coded using the World Health Organization (WHO) Drug Global coding dictionary (September 2021 version).

## Data Availability

All data produced in the present study are available upon reasonable request to the authors

## ACKNOWLEDGMENTS

This study was funded by Atea Pharmaceuticals, Inc and F Hoffmann-LaRoche Ltd. Medical writing support was provided by Samantha Brick, Elements Communications Ltd, UK, and funded by Atea Pharmaceuticals, Inc.

Xiao-Jian Zhou, Arantxa Horga, Maureen Montrond, Keith Pietropaolo, Bruce Belanger, and Janet Hammond are employees of and may own stock in Atea Pharmaceuticals, Inc, Boston, MA, USA; Adeep Puri is an employee of Hammersmith Medicines Research Ltd, London, UK, which was contracted to help perform this research; Lee Winchester and Courtney V. Fletcher are employees of the University of Nebraska Medical Center, Omaha, NE, USA, and were contracted to help perform this research.

